# Visual Fidelity–Driven Quality Assessment of Medical Image Translation

**DOI:** 10.64898/2026.03.18.26348721

**Authors:** Žiga Bizjak, Jan Žagar, Žiga Špiclin

## Abstract

Automated and reliable image quality assessment (IQA) is essential for safe use of medical image synthesis in critical applications like adaptive radiotherapy, treatment planning, or missing-modality reconstruction, where unnoticed generative artifacts may adversely affect outcomes. We evaluated image-to-image translation quality by coupling large-scale expert visual quality assessment with explainable automated IQA modeling. Adversarial diffusion-based framework, SynDiff, was applied to four cross-modality synthesis tasks, including three inter-MR and a CBCT-to-CT translation. Using four-fold cross-validation, ten reference-based and eight no-reference IQA metrics were computed for all synthesized images. Visual IQA ratings were independently collected from thirteen expert raters using predetermined protocol and specialized image viewer enabling blinded, randomized six-point Likert scoring. Auto-Sklearn was employed to learn ensemble regression models mapping IQA metrics to visual consensus ratings, with separate models trained on reference-based and no-reference metrics. The models closely reproduced distribution and ordering of expert ratings, typically within ± 0.5 Likert points. Reference-based models achieved higher agreement with visual ratings than no-reference models (*R*^2^ 0.75 vs. 0.59, resp.), although the latter remained unbiased and informative. Explainability analyses highlighted structure- and contrast-sensitive metrics as key predictors. Overall, the results demonstrate that ensemble regression models can provide transparent, scalable, and clinically meaningful quality control for generative medical imaging.

## Introduction

Medical imaging increasingly relies on complementary modalities to capture diverse anatomical and pathological information; however, in routine clinical practice, acquiring multiple scans is often constrained by cost, acquisition time, and/or radiation exposure^1,2^. Medical image-to-image translation has therefore emerged as a key computational approach to address these limitations by synthesizing missing modalities or enhancing existing images using generative artificial intelligence^3^. Such techniques enable clinically relevant high-stakes applications including cross-modality synthesis for radiotherapy planning^4^, low-dose to standard-dose image enhancement to reduce patient risk, missing modality recovery and data augmentation for training robust diagnostic models, to anomaly detection and image-guided interventions^5^. The model architectures advanced from traditional generative adversarial networks toward more stable and expressive ones, such as diffusion models and transformer-based frameworks, resulting in substantial improvements in image fidelity and anatomical consistency^6,7^. However, a critical barrier to deploying generative image-to-image translation models in high-stakes clinical workflows is the lack of reliable automated image quality assessment (IQA) for synthesized images.

In medical imaging the IQA has traditionally relied on visual inspection, where trained clinicians or engineers judge image fidelity, anatomical plausibility, and the presence of artifacts based on domain-specific experience^8^. While human visual assessment remains the gold standard, it is subjective, time-consuming, and difficult to scale; for instance, a review found that clinical software validation involved a median of four health-care professionals, potentially limiting generalizability^9^. This clearly motivates the use of quantitative IQA metrics as objective surrogates for perceptual quality.

Quantitative IQA metrics can be broadly divided into *reference-based* and *no-reference*^10^. Reference-based metrics require paired data with an available ground-truth image of the same modality and include measures such as peak signal-to-noise ratio (PSNR), structural similarity index (SSIM), and multi-scale SSIM, which quantify pixel-wise fidelity, contrast, and structural agreement^10,11^. In contrast, no-reference metrics operate without a reference image and instead rely on statistical or perceptual cues intrinsic to the image, such as the Natural Image Quality Evaluator (NIQE)^12^, entropy-based measures, or blur-sensitive indices like cumulative probability of blur detection (CPBD)^13,14^. This distinction has direct implications for automated IQA: reference-based approaches are typically more sensitive and accurate but limited to paired datasets, whereas no-reference metrics enable deployment in unpaired or real-world clinical scenarios where ground truth is unavailable, albeit with reduced specificity. A study^15^ established a benchmark dataset to evaluate the capacity of 12 reference-based and 11 no-reference IQA metrics in detecting MR-specific distortions in synthesized brain scans. Findings indicate that traditional metrics are unreliable for validating the synthetic scans, highlighting the need for specialized, multi-tiered evaluation to ensure clinical safety. Beyond generic IQA, task-driven metrics derived from downstream applications, such as segmentation accuracy, registration error, or dosimetric deviation in radiotherapy, have also been proposed^16^, but these are inherently task- and model-dependent and may limit the generalizability of automated IQA frameworks across imaging modalities and clinical use cases.

Research on quantitative IQA relation to human perception has primarily focused on natural image databases like Tampere Image Database^17^ and LIVE Image Quality Assessment Database^18,19^. However, findings therein cannot be directly transferred to the medical domain without task-specific revalidation. For instance, standard reference-based metrics such as PSNR and SSIM often showed a poor or inconsistent correlation with human expert ratings in medical contexts, particularly because they were insensitive to clinically critical, localized anatomical details. Recent comparative studies utilizing expert radiologists have identified HaarPSI^20^ as a consistently high-performing metric across various medical data sets, including chest X-ray and photoacoustic imaging, whereas SSIM ranked among the worst-performing metrics for these tasks. Among no-reference metrics, general-purpose metric like NIQE^12^ struggled to accurately identify quality decreases in specialized medical data, such as accelerated MRI reconstructions. These gaps underscore the urgent need for human evaluator studies and standardized benchmark datasets and tools to systematically assess image-to-image translation performance. In particular, to capture clinically relevant factors such as pathological lesion (e.g., tumor) visibility and modality-specific artifacts (e.g., ghosting or bias fields), the scientific consensus highlights the necessity of developing tailored medical IQA metrics or multi-tier evaluation approaches, complemented by expert-annotated medical databases^21^.

### Contribution

In this study, we make the following key contributions: **(i)** we provide a large-scale evaluation of medical image-to-image translation quality by combining expert visual assessment with explainable automated IQA modeling; **(ii)** we apply an adversarial diffusion-based framework, SynDiff, across four cross-modality synthesis tasks, including inter-MR and CBCT-to-CT translations; **(iii)** we systematically compute both reference-based and no-reference IQA metrics and map them to expert consensus ratings using Auto-Sklearn ensemble regressors; **(iv)** we demonstrate that the models closely reproduce human ratings, with reference-based metrics achieving higher agreement while no-reference metrics also remain informative; and **(v)** we identify the most influential metrics driving predictions, providing insight into the structural and contrast-based factors critical for clinically meaningful quality control. Together, these contributions establish a transparent, scalable framework (Figure 1) for automated IQA in generative medical imaging.

**Figure 1.**
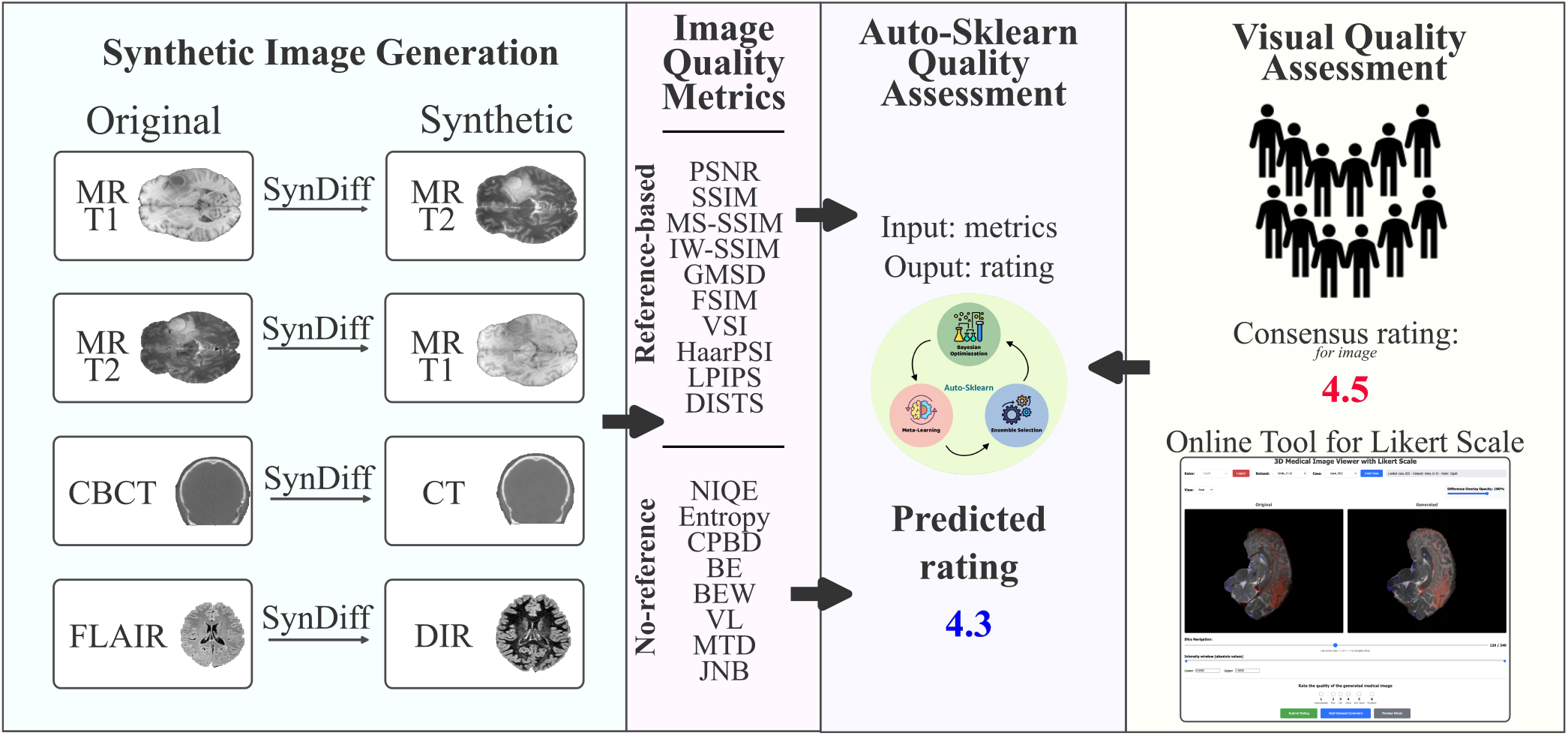
Study design overview: in four image-to-image translation tasks, carried out by training the SynDiff model^22^, the synthesized images were quantitatively assessed using reference-based and no-reference metrics and blinded Likert-based visual assessment by 13 raters, followed by mapping and analysing the metric values versus the consensus visual ratings.

## Results

### Visual Quality Assessment

Consensus visual image quality ratings spanned from 1.0 to 5.07 on the 1–6 Likert scale, indicating that raters effectively exploited the full range of the assessment scale. The median consensus ratings were 3.2, 3.7, 2.6, and 4.2 for the `T1` → `T2`, `T2` → `T1`, `CBCT` → `CT`, and `FLAIR` → `DIR` image-to-image translation tasks, respectively. These results clearly indicate that the `CBCT` → `CT` translation task was the most challenging, which can plausibly be attributed to the greater heterogeneity of CBCT acquisitions, including variability across imaging systems and substantial differences in intensity distributions, potentially arising from the lack of standardized Hounsfield unit calibration. On the other hand, the scans for the `FLAIR` → `DIR` task dataset originated from same scanner and the corresponding SynDiff model produced only good- to high-quality synthesized DIR scans (range 3–6 on the Likert scale). Figure 2 shows a representative selection of cases across the Likert scale range for the four datasets and tasks.

**Figure 2.**
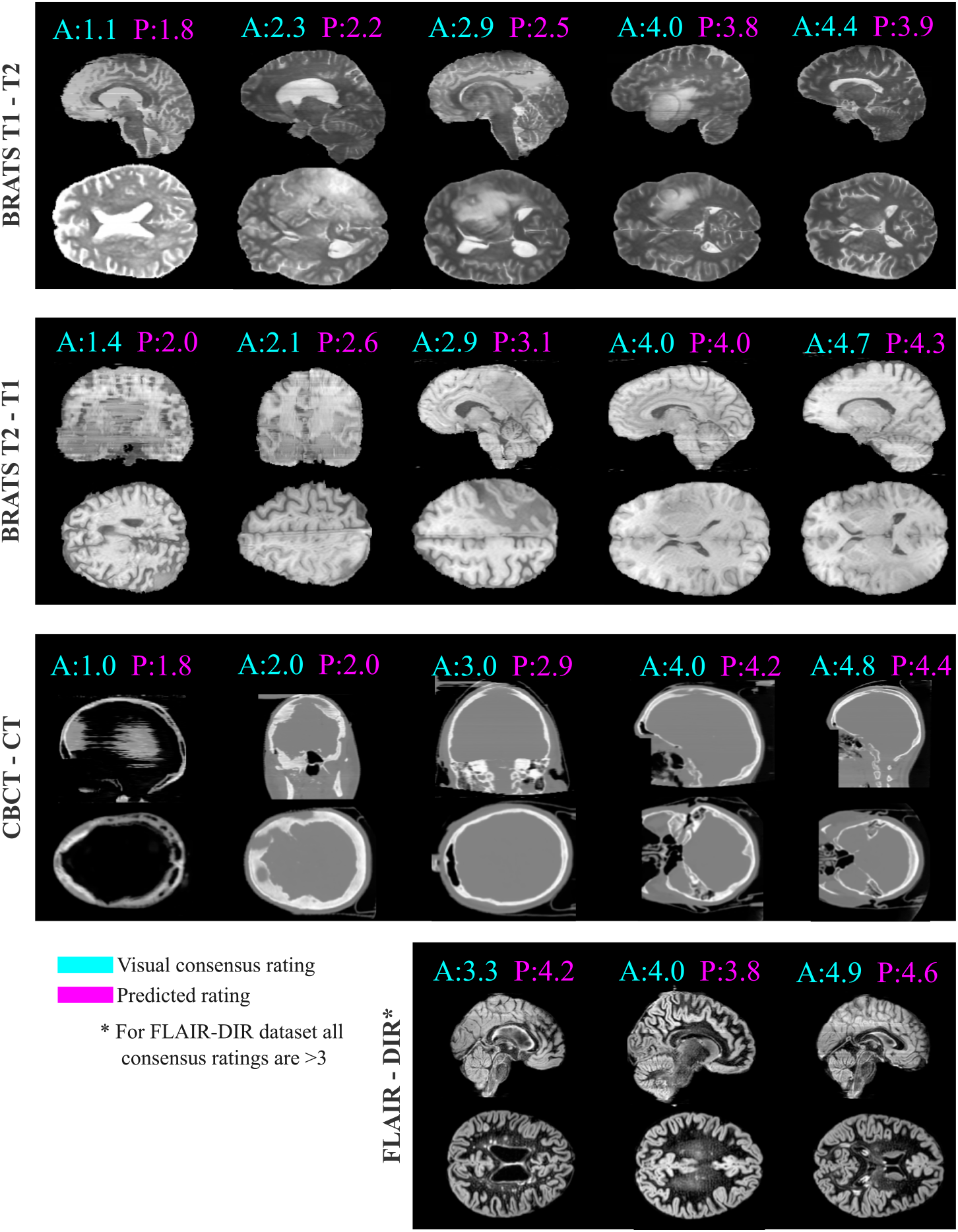
Qualitative comparison of (A) visual consensus versus (P) model-predicted ratings across the four tasks. The regression models successfully replicated visual consensus ratings across a wide spectrum of the 6-point Likert scale, ranging from synthetic scans with severe artifacts to high-fidelity ones in the `T1`→`T2`, `T2`→`T1` and `CBCT`→`CT` tasks. In `FLAIR`→`DIR` task, the model correctly reproduced good to high ratings consistently with human raters.

### Automated Quality Assessment

Two sets of models predicting the consensus visual ratings were trained using Auto-Sklearn, using reference-based and no-reference IQA metric values as input. The performance of the automated QA models was evaluated using 4-fold cross-validation across *N* = 287 cases.

The resulting models, regardless of the input IQA metrics and particular fold, were a weighted `VotingRegressor` ensemble comprising several distinct pipelines. For instance, for the model using reference-based IQA metrics a best validation *R*^2^ of 0.7475 was achieved. The primary predictive strength was derived from five AdaBoost variants using Decision Tree base learners with weights totaling approximately 0.60. These learners ranged from shallow depth-3 trees to deep depth-9 trees, allowing the model to capture both broad quality trends and specific, localized generative artifacts. Additionally, with a weight of 0.2267, a significant portion of the prediction logic relied on Support Vector Regression (SVR) (*C* ≈ 2.1*e*3, *γ* ≈ 0.0152). A Multi-Layer Perceptron (MLP) component with three hidden layers of 19 units each contributed a weight of 0.12, providing a functional alternative to the tree-based logic, while a minor contribution from an ExtraTreesRegressor (weight: 0.0067) added a final layer of variance reduction to the ensemble.

The model-predicted image quality ratings well reproduced the actual ratings’ distribution by spanning from 1.60 to 4.73. Qualitative comparison of the actual visual consensus (*A*) with model-predicted ratings (*P*) is shown in Figure 2. In the BraTS2020 tasks `T1` → `T2` and `T2` → `T1`, the models accurately captured the transition from heavily artifacted, low-rated scans to high-fidelity images. Similarly, in the `CBCT` → `CT` task, the models correctly identified substantial geometric distortions and noise, assigning scores near 1.0, while rating high-fidelity synthetic images near 5.0. In the `FLAIR` → `DIR` task the models exhibited high ratings (*>* 3.0) consistent with the visual consensus assessment. The predicted values approximated well the human raters’ consensus values, typically within a 0.5-point margin on the 6-point Likert scale.

Overall agreement statistics and distribution data are summarized in Table 1. The model using reference-based IQA metrics achieved a mean *R*^2^ of 0.752 and a mean absolute error (MAE) of 0.374, while the model using no-reference IQA metrics had comparatively lower agreement according to lower mean *R*^2^ of 0.589 and higher MAE at 0.478.

**Table 1.** Summary of Automated QA Model Performance and Distribution Statistics.

| Category | Metric | Reference-based | No-reference |
| --- | --- | --- | --- |
| Agreement Statistics | $R^2$ [Min, Max] | 0.752 [0.683, 0.791] | 0.589 [0.441, 0.659] |
|  | MAE [Min, Max] | 0.374 [0.351, 0.401] | 0.478 [0.431, 0.518] |
| Rating Distribution | Actual Mean ± SD | $2.97 \pm 0.99$ | $2.97 \pm 0.99$ |
| | Predicted Mean ± SD | $2.98 \pm 0.87$ | $2.99 \pm 0.80$ |
| | Overall $p$-value* | 0.8596 | 0.641098 |
| | Per-Dataset $p$-value* Range | 0.413 – 0.749 | 0.554 – 0.696 |
|  | Median Bias [95% Bootstrap CI] | -0.018 [−0.090, 0.051] | 0.0232 [−0.0517, 0.0903] |
|  | Per-Dataset Median Bias Range | [−0.173, 0.030] | [−0.2927, 0.3167] |

MAE: Mean Absolute Error; SD: Standard Deviation; CI: Confidence Interval; \*Based on Wilcoxon signed-rank test.

The predicted ratings (*µ* = 2.98 ± 0.87 and 2.99 ± 0.80, for reference-based and no-reference IQA metrics, resp.) closely followed the visual consensus ratings (*µ* = 2.97 ± 0.99). According to high *p*-values and negligible median bias there was no significant differences in the ratings’ distributions. These results were consistent across all sub-tasks (`T1` → `T2`, `T2` → `T1`, `CBCT` → `CT`, `FLAIR` → `DIR`), with non-significant per-dataset *p*-values and no systematic over- or under-estimation of quality regardless of the imaging modality according to small median bias values.

The relationship between actual visual consensus ratings and prediction output by the model is visualized in Figures 3ab, where each imaging dataset is represented by a distinct color. The linear fit for model using reference-based IQA metrics (*y* = 0.76*x* + 0.72) follows the ideal diagonal across all modalities rather well, with most data points falling within the shaded ± 1 standard deviation interval. This demonstrates that the model remained consistent across different data sources and rarely deviated from visual consensus assessment by more than one point on the Likert scale. In comparison, the linear fit for model using no-reference IQA metrics (*y* = 0.62*x* + 1.14) was more systematically biased and with higher random error as also indicated by the generally lower *R*^2^ and higher MAE values (Table 1).

**Figure 3.**
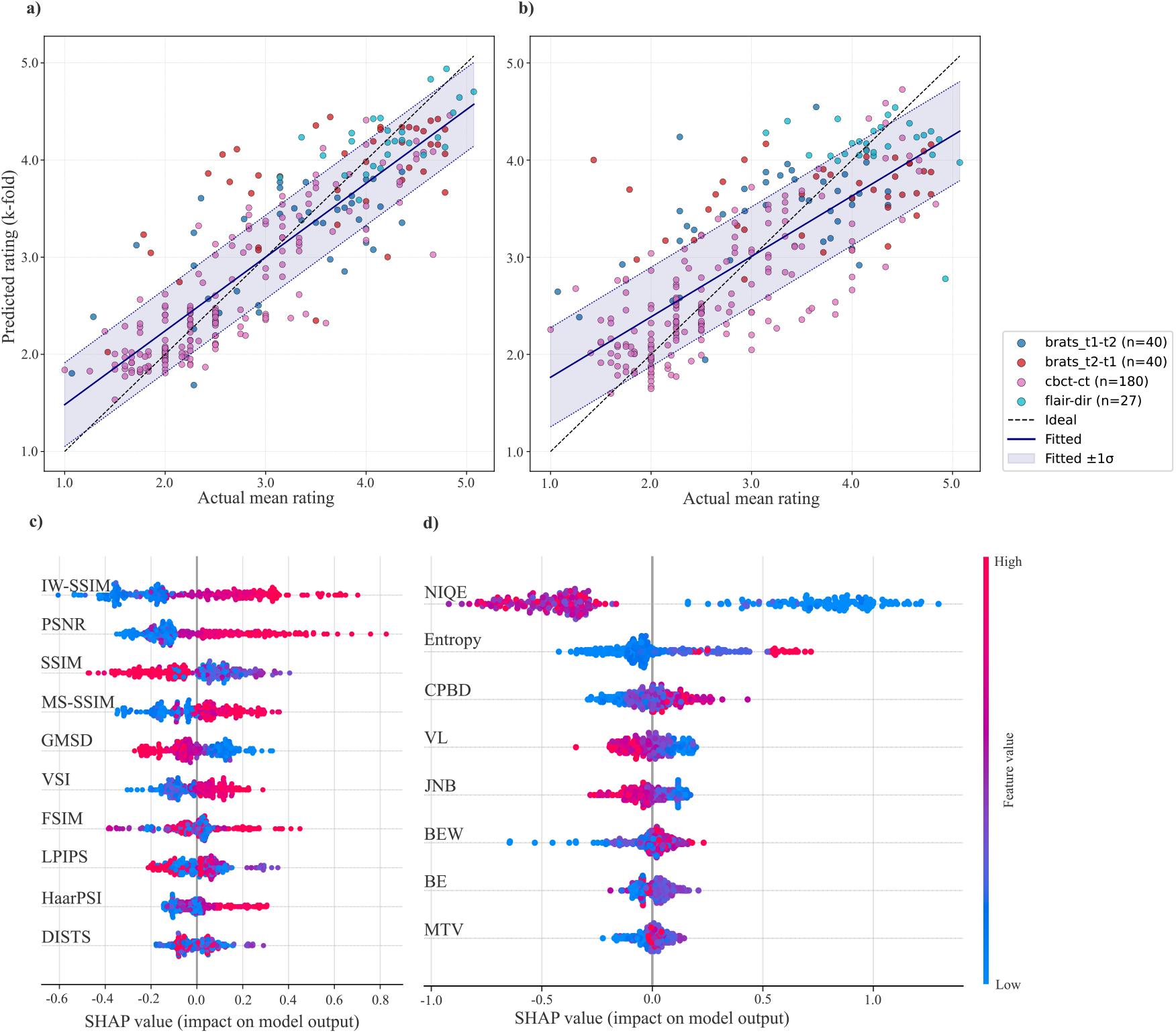
Scatter plots of actual mean human ratings versus k-fold predicted ratings (*N* = 287), with shaded ± 1*σ* confidence interval, for (a) reference-based and (b) no-reference metrics. Respective SHAP summary plots ranking the metrics are shown in (c) and (d). The metric values (low to high) are represented by color, where positive SHAP values indicate an increase in the predicted quality rating.

#### Explainability Analysis

Impact of the IQA metrics on the model output is provided in the SHAP summary plots in Figures 3cd. Among the reference-based IQA metrics the SHAP analysis identified IW-SSIM, PSNR, and SSIM as the most influential metrics. The predicted quality rating was consistently proportional to the magnitude of the IW-SSIM and PSNR metrics. Among the no-reference IQA metrics the SHAP analysis identified NIQE, entropy and CPBD as the most impactful metrics. The small magnitude of NIQE metric was clearly associated with high predicted quality rating. Such obvious dichotomy between IQA metrics and positive model output confirms that the regressor has successfully learned to replicate the ranking criteria used by human experts and that visual quality assessment automation is feasible based on certain IQA metrics.

The stability of IQA metrics’ importance across the cross-validation folds is illustrated in Figure 4. Among the reference-based IQA metrics, the IW-SSIM emerged as the most robust predictor, consistently maintaining a top-two rank across all folds. While other structural metrics such as MS-SSIM and FSIM frequently rank among highest, they exhibit periodic fluctuations in rank depending on the specific training fold partition – most notably the drop in MS-SSIM rank in Fold 2 and FSIM in Fold 3. Despite these minor variations, the ensemble model primarily relied on a consistent subset of contrast- and structure-sensitive IQA metrics to predict QA ratings.

**Figure 4.**
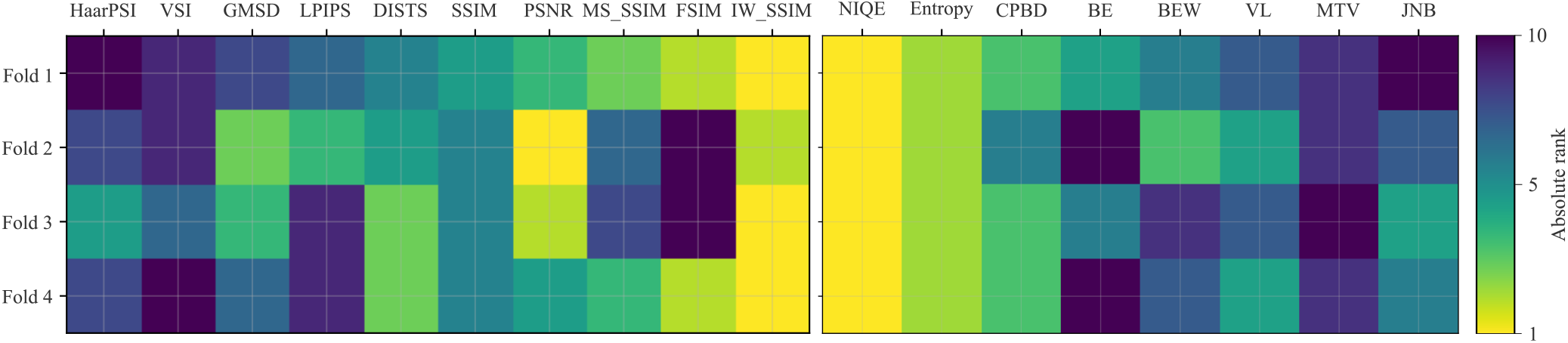
Absolute rank (1 = strongest) of each IQA metrics across the four cross-validation folds (*rows*) for models using reference-based (*left*) and no-reference metrics (*right*). *Brighter tiles* indicate more informative metrics.

Among the no-reference metrics, the intensity-statistics–based NIQE consistently exhibited the highest importance, followed by the entropy metric, indicating that global intensity distributions and structural information are crucial in predicting quality assessment ratings. Metrics specifically sensitive to blurring artifacts, namely CPBD, BE, BEW, and VL, ranked third to sixth in importance and provided complementary contributions by capturing loss of sharpness and over-smoothing effects.

#### Sensitivity Analysis

Figure 5 shows aggregated partial dependence plots (PDP) in order to evaluate the marginal effect of individual IQA metrics on the model’s predictions. For regression model based on reference-based IQA metrics, the metrics focused on image structure, such as IW-SSIM and MS-SSIM, and contrast-sensitive PSNR, exhibited a clear monotonic proportional relationship: as the metric value increased, the predicted QC rating increased accordingly. Conversely, distortion-based metrics like GMSD, LPIPS, and DISTS showed an inverse relationship, where higher values (indicating greater image degradation) were associated with the decrease in the predicted rating. Interestingly, the SSIM did not exhibit a monotonic relationship with the visual consensus score; this non-monotonic behavior arises from the bounded and saturating nature of SSIM and its reliance on local second-order statistics. While SSIM correlates with visual quality for large structural discrepancies, it becomes insensitive to subtle yet clinically relevant artifacts at intermediate values and may over-reward excessive smoothing or locally consistent hallucinations at high values, leading to an inverse relationship with expert visual assessments.

**Figure 5.**
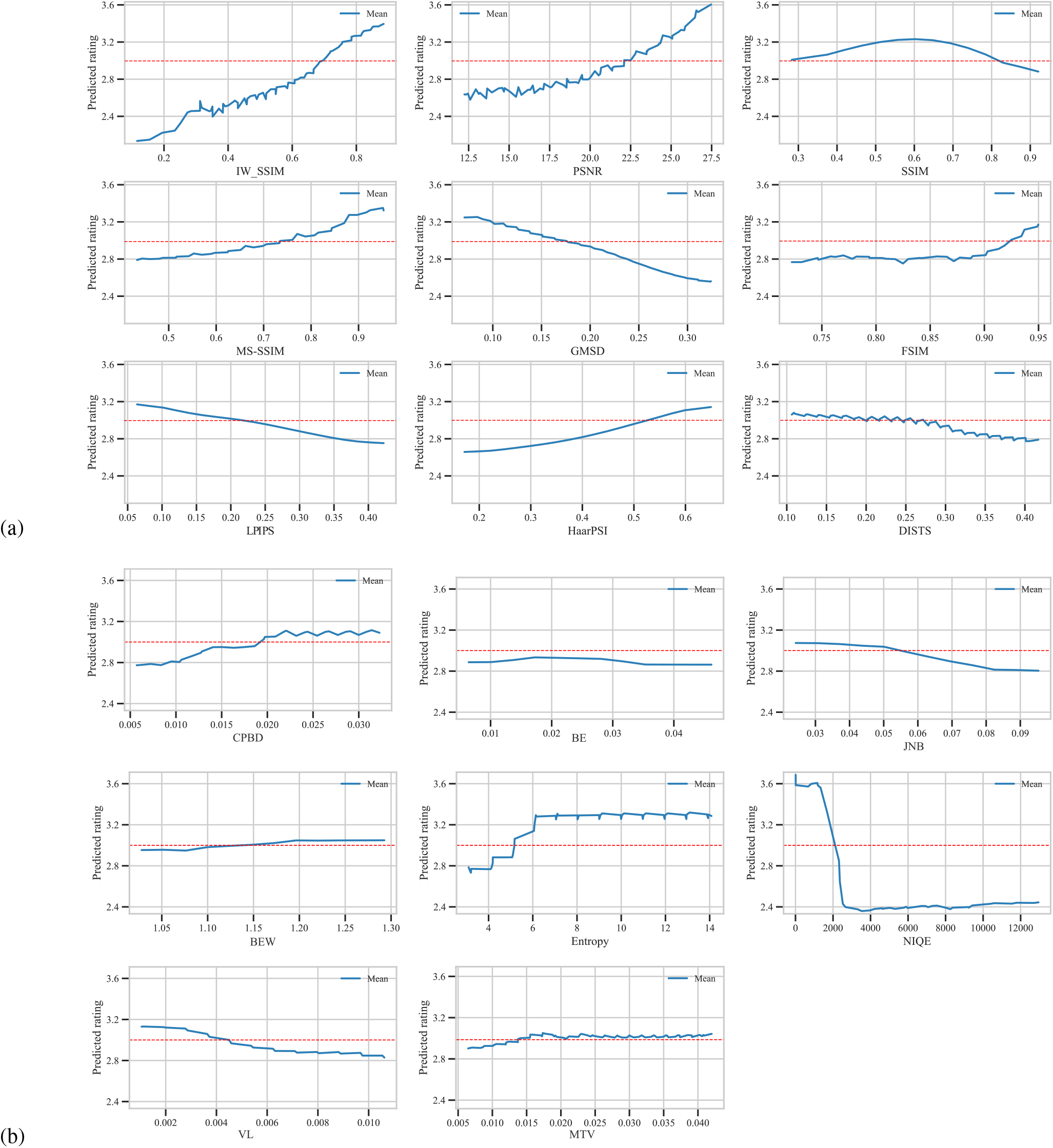
Partial dependence plots aggregated across four test folds after five-point rolling smoothing for prediction on regression models using (a) reference-based and (b) no-reference IQA metrics. Each panel traces *in blue line* the mean predicted QA rating with respect to specific metric value, while *dashed red line* indicates the mean consensus visual IQA rating.

Analysis of the model using no-reference IQA metrics showed the NIQE exhibited the strongest and most structured effect: with low NIQE values corresponding to a steep increase in predicted ratings, whereas a broad range of high NIQE values was consistently associated with uniformly low ratings.

In contrast, CPBD and entropy demonstrated approximately proportional relationships with predicted ratings, indicating that edge sharpness and information content contribute positively to perceived image fidelity. However, both metrics became largely insensitive at higher magnitudes, reflecting known limitations whereby excessive sharpness or entropy may arise from noise or non-physical textures that do not translate into improved perceptual quality. Metrics explicitly designed to capture blur magnitude, such as BE, BEW, and MJV showed minimal influence on model predictions, suggesting that these measures may be too localized or narrow in scope to explain the broader spectrum of generative artifacts encountered in diffusion-based synthesis. Finally, JNB and VL exhibited weak inverse relationships with predicted ratings, consistent with their sensitivity to high-frequency content; however, the low effect size and instability across the value range indicate that these associations may reflect secondary or potentially spurious effects rather than dominant perceptual drivers.

## Discussion

This study addressed the development, evaluation, and analysis of automated IQA models based on ensemble regressors, relating reference-based and no-reference IQA metrics to blinded 6-point Likert ratings aggregated across 13 raters. Such models are essential for validating generative image-to-image methods in medical imaging, where quality must be assessed with respect to clinically relevant visual fidelity. The inclusion of diverse datasets (BraTS, CBCT-CT, and FLAIR-DIR) ensured heterogeneous imaging conditions, reducing the risk of overfitting to a single modality or tissue type.

The full range of human IQA ratings was well represented across three of the four datasets (Figure 3ab). Although the maximum possible rating was 6.0, no image achieved perfect consensus; the highest mean rating was 5.07. This ceiling effect indicates that even high-quality synthesized images contain subtle, expert-detectable deviations, reflecting the rigor of the evaluation protocol requiring artifact localization and justification.

Images were generated using the SynDiff model, a 2D diffusion-based architecture. While efficient, such models lack full 3D context and may introduce artifacts such as inter-slice discontinuities or checkerboard patterns. By explicitly training raters to identify these artifacts, the resulting regression models capture perceptually relevant degradations often missed by conventional metrics, enabling scalable yet human-aligned IQA.

Quantitatively, regression from IQA metrics to human ratings proved effective, particularly for reference-based metrics (*R*^2^ = 0.752, MAE = 0.374), with somewhat reduced but still meaningful performance for no-reference metrics (*R*^2^ = 0.59). The ensemble approach demonstrated robustness across folds and datasets, consistently relying on structure- and contrast-sensitive metrics such as IW-SSIM, MS-SSIM, and NIQE. Partial dependence analysis confirmed that model predictions follow physically meaningful trends, with structural fidelity positively associated with perceived quality. The prominence of IW-SSIM further suggests that information-weighting aligns well with human prioritization of anatomically relevant regions.

These findings are broadly consistent with the systematic analysis by Dohmen et al.^**?**^, who demonstrated that no single IQA metric sufficiently captures perceptual quality and that combinations of complementary metrics are required. Both studies highlight limitations of commonly used measures such as SSIM and PSNR, including sensitivity to intensity scaling, spatial misalignment, and reduced sensitivity to blurring or over-smoothing. Our observation of non-monotonic SSIM behavior further supports these known shortcomings.

However, discrepancies are also evident. Dohmen et al. report instability of certain no-reference metrics, particularly NIQE, whereas in our study NIQE emerged as a strong predictor of perceived quality. This difference likely arises from experimental design: their controlled distortion framework on homogeneous MRI data contrasts with our evaluation on real generative outputs across multiple modalities. In this context, NIQE may act as a proxy for global statistical deviations rather than specific distortions. Similarly, while PSNR is discouraged in isolation, our results indicate that it retains value within an ensemble, suggesting that metric limitations can be mitigated through complementary integration.

The key contribution of this work is the transition from controlled, modality-specific metric analysis to a visual perception grounded, multi-task evaluation framework anchored in large-scale human assessment. By explicitly modeling the relationship between IQA metrics and human consensus ratings, and leveraging explainable methods such as SHAP, we provide insight into how objective measures translate to perceived quality in practice. This also enhances interpretability and supports the development of trustworthy AI systems, which is critical for clinical adoption.

From a practical perspective, such automated IQA models enable scalable quality control of generative models, including potential real-time assessment during training or deployment, and rejection of clinically inadequate outputs. However, robustness to domain shift (e.g., scanner variability or acquisition protocols) remains an important consideration, motivating future work on domain-adaptive calibration.

This study has several limitations. It is restricted to a single generative framework (SynDiff), and it remains unclear how well the learned relationships generalize to other architectures (e.g., GANs^23^, diffusion models^24,25^ and transformers^26^). Additionally, the datasets were limited to brain imaging, and ratings were obtained from a relatively homogeneous group of expert raters. To address this, we will release the annotation tools and protocols to facilitate broader, multi-center validation studies. Future work should evaluate generalization across modalities, tasks, and institutions, and explore task-specific or adaptive IQA models.

In conclusion, this work demonstrates that automated IQA models can reliably approximate expert visual assessment across diverse medical image synthesis tasks. By bridging objective metrics and human perception, the proposed framework enables transparent, scalable, and clinically meaningful quality control. The open-source release of tools and models will further promote standardization and reproducibility, ultimately supporting safer and more reliable integration of generative AI into clinical practice.

## Materials and Methods

### Datasets

Dataset with a total of *N* = 287 subjects, each with paired image modality cases, was sourced and curated from three cross-modality datasets. Two were MR image datasets, first a `t1-t2` dataset comprised of 80 pairs of T1- and T2-weighted (T1w and T2w) scans from the BraTS2020 Brain Tumor Segmentation Challenge^27–29^, while the second a private `flair-dir` dataset that included 27 multiple sclerosis subjects, each imaged with T1-weighted, FLAIR and DIR scans^30^ (approval no. 0120-660/2015-2). All images were spatially normalized using ANTsPy with mutual information–based affine registration^31^. For each subject, all scans were first registered to the corresponding T1-weighted (T1w) image, after which the subject’s T1w scan was registered to the T1w template of the MNI2009c brain atlas^32^. All multi-modal brain MR scans were skull-stripped based on 3-pixel dilated MNI2009c atlas brain mask and normalized by windowing intensities using the white stripe normalization method^33^.

The third dataset was `cbct-ct` and consisted of 180 CBCT-CT brain images pairs from the SynthRAD2023 Task2 training dataset^34^, which were already rigidly co-registered. These images were linearly windowed between -1000 and 1500 HU, which captured over 99.8% of the intensity distribution.

### Image-to-Image Translation

For image-to-image translation we used the SynDiff model^22^, due to code availability^1^ and its robust performance. It is an unsupervised adversarially trained conditional diffusion process for synthesizing images across modalities.

Models were trained independently for each of four image translation tasks. For the `t1-t2` dataset, two separate models were trained, namely for the tasks of `T1` → `T2` and `T2` → `T1` translation, while for the `flair-dir` and `cbct-ct` datasets, the `FLAIR` → `DIR` and `CBCT` → `CT` translation models were trained, respectively. Co-registered volumetric images were sampled as 2D axial slice stacks, with each slice resized to 256 *×* 256 pixels and min–max intensity normalized to the range [0, 1]. Using all 287 subject datasets, model training was performed in a four-fold cross-validation scheme.

In each cross-validation split, the held-out test fold was used exclusively for inference, and for each test subject the corresponding reference image of the target modality was available. This setup enabled robust and unbiased comparative performance evaluation of the synthesized images against ground-truth references.

### Visual Quality Assessment

To streamline the collection of high-quality ground truth data, we developed a specialized image quality assessment application that enables single-blinded, randomized Likert-scale visual evaluation of (medical) image volumes. This section describes the Medical Image Viewer with Likert Scale, optimized for efficient and standardized visual assessment, as well as the rater recruitment, annotation protocol definition, rater training, and quality assurance procedures used to ensure consistent and reliable expert ratings.

#### Medical Image Viewer with Likert Scale

The visual quality assessment application was implemented in Python 3.9 using the Flask framework and Flask-SocketIO for real-time communication, providing a responsive environment for effective image evaluation. The system allows to load and display standard medical image formats (DICOM, NIfTI, etc.). The viewer provides a side-by-side slice-wise comparison of 3D volumes. It provides standardized radiological capabilities, including multi-planar reconstruction (axial, sagittal, and coronal views) and dynamic intensity windowing (level and width adjustment). The rater view is shown in Figure 6.

**Figure 6.**
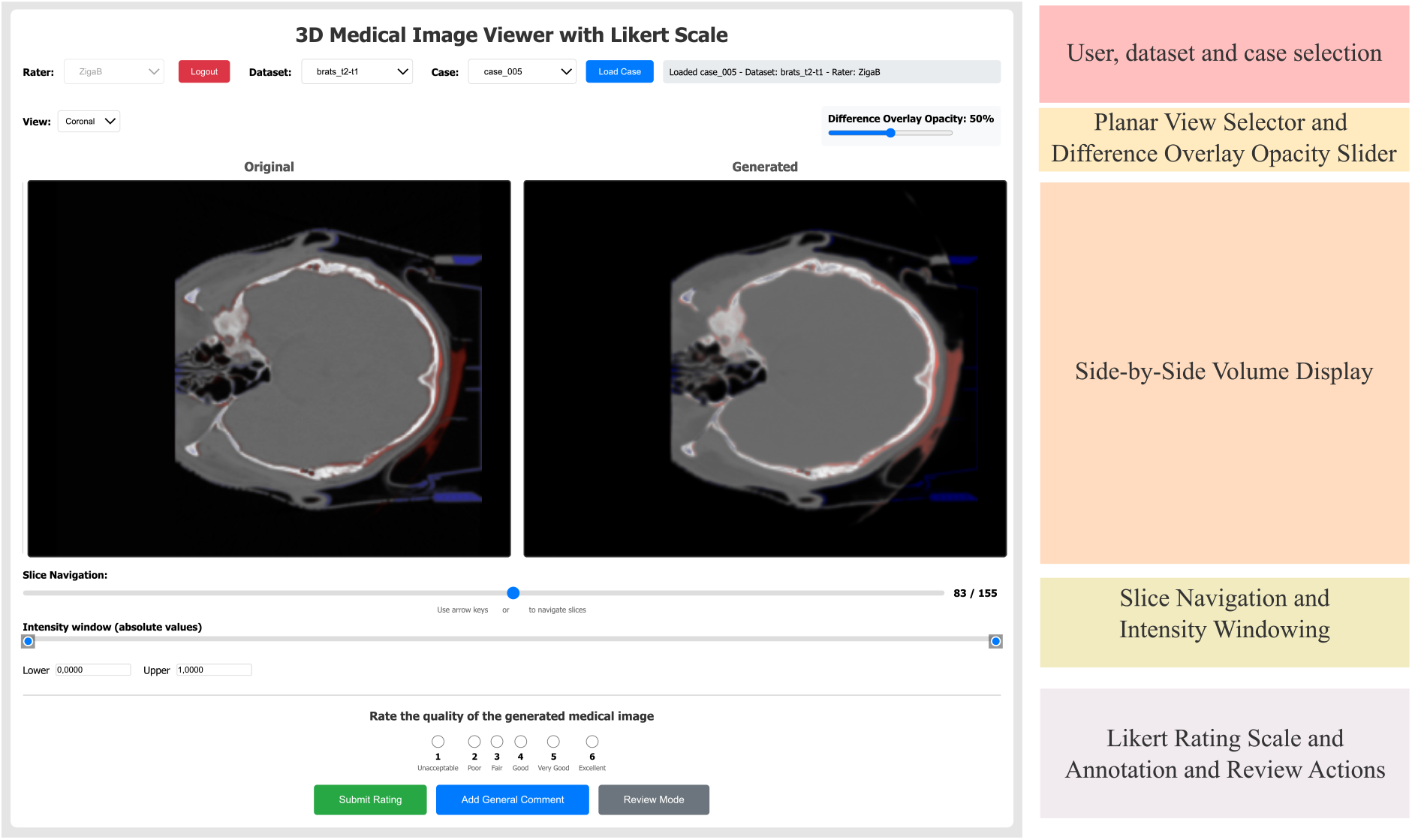
Layout of the graphical user interface of the Medical Image Viewer with Likert Scale used for visual quality assessment. All actions were associated to keyboard shortcuts to expedite the assessment process.

Since the aim was to compare the original and synthesized volumes, a key technical feature to better highlight differences is the color-coded overlay of pixel-level discrepancies – using red for positive differences (synthesized *>* original) and blue for negative – with adjustable opacity to assist raters in identifying localized artifacts or structural hallucinations.

To facilitate granular feedback, the interface allows raters to place point-based annotations directly on the image to highlight specific generative defects, accompanied by detailed text comments. Likert quality rating is achieved using radio buttons, while raters must provide a general comment for justifying their rating.

The application has two access levels: `administrator` and `user`. The access level is strictly controlled through a password-protected authentication system. The `administrative` level provides a centralized dashboard for reviewing all submitted ratings and annotations across the entire rater pool, ensuring the integrity of the ground truth data, and enables setting the annotation protocol, i.e. data sources, case name and order randomization, required fields, etc. The `user` level executes the predefined annotation protocol and provides raters with an intuitive interface for blinded image evaluation, including anonymized and randomized case selection that enables repeated assessments while minimizing memorization effects. Raters interact with the system exclusively through the active protocol, ensuring consistency across evaluations, and all core actions—such as slice navigation, view switching, annotation placement, opacity adjustment, and Likert score selection—are mapped to keyboard shortcuts to streamline interaction and expedite the annotation and rating process.

To ensure ease of use and reproducibility, the entire application is dockerized, enabling rapid, consistent deployment across different platforms. Full source code and pre-configured Docker containers will be made publicly available on GitHub^2^ to support community efforts in standardizing generative AI validation.

#### Raters, Annotation Protocol and Quality Assurance

A team of 13 raters was recruited to perform blinded evaluations of the synthesized volumes. Additionally, two senior researchers in medical image analysis acted as supervisors. The raters were master students enrolled in final-year biomedical engineering program with relevant 12-months prior coursework in biomedical image processing and medical image analysis, providing solid experience in image annotation and image processing algorithm design and validation.

Using the visual quality assessment application in `user` level all raters assessed visual image fidelity and clinical utility on a Likert scale according to the following discrete six-level scale:

| *Rating* | *Criterion description* |
| --- | --- |
| 1 – Unacceptable | Severe artifacts or distortions; image is not interpretable and has no clinical or analytical value. |
| 2 – Poor | Major artifacts or unrealistic appearance; interpretation is highly limited and clinically unreliable. |
| 3 – Fair | Noticeable artifacts or loss of detail; image partially interpretable but of limited clinical usefulness. |
| 4 – Good | Minor artifacts present; image is largely realistic and clinically usable with some limitations. |
| 5 – Very Good | High visual fidelity with subtle imperfections; image clinically useful and comparable to real data. |
| 6 – Excellent | Indistinguishable from real images; no visible artifacts, fully suitable for clinical and analytical use. |

Prior to the *evaluation phase* all 13 raters underwent a standardized training session by the two supervisors to ensure high intra-rater reliability and relevance of the assessments. This training familiarized the raters with the application’s graphical interface and established a shared criteria for identifying generative artifacts, such as anatomical hallucinations, blurred boundaries, and inter-slice discontinuities inherent to the use 2D diffusion model.

For traceability and quality assurance the rating process required the raters to justify each assigned Likert score by providing either a localized point-based annotation (marking the exact coordinates of an artifact) or a general text comment describing the rationale for the assigned grade. This mandatory requirement ensured that the ratings were grounded in observable evidence rather than subjective impressions.

Following the completion of the evaluation phase performed by all 13 raters, the supervisors performed a secondary review of the data, conducted using the `administrator` level in application. During this phase, the obtained values were analyzed to identify statistical outliers or inconsistent annotations. Ratings that deviated significantly (i.e. 3 points on the Likert scale) from the median value without sufficient justification were removed to maintain the integrity of the training set. Finally, the remaining expert scores were aggregated to compute a mean Likert rating for each case.

### Image Quality Metrics

Ten reference-based and eight no-reference image quality metrics were computed as summarized in Table 2. Synthesized images generated on the test fold had corresponding reference images of the same modality, based on which the reference-based image quality metrics were computed. Multiple metrics were evaluated since individual metrics might have specific “blind spots.” For example, while SSIM measures structural coherence, it often misses the non-physical textures or artifacts common in diffusion-based models. Metrics like LPIPS or HaarPSI are better suited to detecting these high-level perceptual errors. On the other hand, the no-reference image quality metrics do not rely on the availability or fidelity of a paired ground-truth image, therefore enabling robust and scalable quality control at inference time. They allow the detection of excessive smoothing (BE, BEW, JNB, CPBD) and noise amplification (entropy, TV, MTV) that may not be penalized by reference-based metrics. Using multiple metrics creates a complementary set of features that may provide a more complete picture of image fidelity compared to individual features. All metric values were evaluated across all test folds for an objective and comprehensive assessment of their association with the visual consensus ratings.

**Table 2.** Metrics used for automated quality assessment of synthesized images.

| Metric | Description | Range | Implementation |
| --- | --- | --- | --- |
| *Reference-based metrics* |  |  |  |
| PSNR<sup>35</sup> | Peak signal-to-noise ratio measuring pixel-wise fidelity | $[0, \infty)$ ↑ | scikit-image<sup>1</sup> |
| SSIM<sup>10</sup> | Structural similarity index comparing luminance, contrast, and structure | $[0, 1]$ ↑ | scikit-image<sup>1</sup> |
| MS-SSIM<sup>11</sup> | Multi-scale extension of SSIM capturing structures at multiple resolutions | $[0, 1]$ ↑ | pytorch-msssim<sup>2</sup> |
| IW-SSIM<sup>36</sup> | Information-weighted SSIM emphasizing perceptually important regions | $[0, 1]$ ↑ | sewar<sup>3</sup> |
| GMSD<sup>37</sup> | Gradient magnitude similarity deviation sensitive to local distortions | $[0, \infty)$ ↓ | sewar<sup>3</sup> |
| FSIM<sup>38</sup> | Feature similarity based on phase congruency and gradient magnitude | $[0, 1]$ ↑ | sewar<sup>3</sup> |
| VSI<sup>39</sup> | Visual saliency-induced index incorporating visual attention modeling | $[0, 1]$ ↑ | sewar<sup>3</sup> |
| HaarPSI<sup>20</sup> | Haar wavelet-based perceptual similarity index | $[0, 1]$ ↑ | sewar<sup>3</sup> |
| LPIPS<sup>40</sup> | Learned perceptual similarity using deep neural network features | $[0, \infty)$ ↓ | lpips<sup>4</sup> |
| DISTS<sup>41</sup> | Deep image structure and texture similarity metric | $[0, 1]$ ↓ | DISTS-pytorch<sup>5</sup> |
| *No reference metrics* |  |  |  |
| NIQE<sup>12</sup> | Natural image quality evaluator based on statistical regularities | $[0, \infty)$ ↓ | medimetrics<sup>6</sup> |
| Entropy<sup>42</sup> | Shannon entropy measuring image information content | $[0, \infty)$ ↑† | medimetrics<sup>6</sup> |
| CPBD<sup>43</sup> | Cumulative probability of blur detection | $[0, 1]$ ↑ | medimetrics<sup>6</sup> |
| BE<sup>44</sup> | Gradient difference before and after additional blurring | $[0, \infty)$ ↑ | medimetrics<sup>6</sup> |
| BEW<sup>45</sup> | Average blurred edge widths | $[0, \infty)$ ↓ | medimetrics<sup>6</sup> |
| VL<sup>46</sup> | Variance of the Laplacian operator (sharpness measure) | $[0, \infty)$ ↑† | medimetrics<sup>6</sup> |
| MTV<sup>47</sup> | Mean total variation (mean L2 gradient magnitude) | $[0, \infty)$ ↓† | medimetrics<sup>6</sup> |
| JNB<sup>48</sup> | Just noticeable blur based on human perception model | $[0, \infty)$ ↓ | medimetrics<sup>6</sup> |

† Might be inflated by increased noise without improving clinical usability of the synthesized image, <sup>1</sup>https://pypi.org/project/scikit-image,
<sup>2</sup>https://pypi.org/project/pytorch-msssim, <sup>3</sup>https://pypi.org/project/sewar, <sup>4</sup>https://pypi.org/project/lpips,
<sup>5</sup>https://pypi.org/project/DISTS-pytorch, <sup>6</sup>https://github.com/bayer-group/mr-image-metrics

### Visual Quality Prediction Modeling and Analysis

The consensus visual image quality ratings and the image-based quality metrics were used to train a regressor capable of predicting the former from the latter. Two distinct models were trained, i.e. first set using reference-based and the second no-reference metrics as input, and their regression performance comparatively evaluated. We utilized the `Auto-Sklearn` framework^49^ – an automated machine learning toolkit that performs holistic architecture search and hyperparameter optimization.

The model optimization process was driven by the *R*^2^ metric, aiming to maximize the explained variance of the consensus visual image quality ratings. We used four-fold cross-validation and prioritized the best-performing ensemble of regressor models across all folds.

For model analysis, we computed agreement statistics such as mean *R*^2^ across all test folds, mean absolute error (MAE), and corresponding min-max ranges. To compare the actual consensus and the predicted ratings we calculated their mean and standard deviations, the p-values of Wilcoxon signed-rank test and median bias. It was defined as the difference between the medians of the two distributions and used to quantify systematic deviations. These performance criteria were computed per- and across datasets. Linear regression model based on root-mean-squared error minimization was evaluated between the predicted and actual values to observe their linear relationship.

Contribution of individual features in the best-performing ensemble regression models was analyzed using SHapley Additive exPlanations (SHAP)^50^, with partial dependence plots illustrating the marginal effects of individual metrics. Analysis was performed separately for models trained on reference-based an no-reference metrics. Each model’s predictions were further validated through qualitative visual inspection of representative synthetic scans in order to confirm sensitivity to relevant artifacts such as blurring and anatomical hallucinations.

## Data Availability

The t1-t2 and t2-t1 datasets were derived from BraTS 2020 challenge dataset that is available at https://www.med.upenn.edu/cbica/brats2020/data.html. The cbct-ct dataset is from the SynthRAD 2023 Task 2 challenge public dataset and is available at https://synthrad2023.grand-challenge.org/. The flair-dir dataset is available from the authors upon reasonable request.

https://synthrad2023.grand-challenge.org/

## Acknowledgments

This study was supported by the Slovenian Research Agency (Core Research Grant No. P2-0232 and Research Grant J2-3059).

## Author contributions statement

Ž.B. and Ž.Š. conceived the study and the experiments, Ž.B. and J.Ž. conducted the experiments, Ž.B., J.Ž. and Ž.Š. analyzed the results. Ž.B. wrote the initial draft, Ž.Š. reviewed and edited the draft. Ž.Š. acquired funding for this study. All authors reviewed the present manuscript and approved it for publication in current form.

## Data availability

The `t1` → `t2` and `t2` → `t1` datasets were derived from BraTS 2020 challenge dataset that is available at https://www.med.upenn.edu/cbica/brats2020/data.html. The `cbct` → `ct` dataset is from the SynthRAD 2023 Task 2 challenge dataset is available at https://synthrad2023.grand-challenge.org/. The `flair` → `dir` dataset is available from the authors upon reasonable request.

## Footnotes

1 [https://github.com/icon-lab/SynDiff

2 GitHub repository: https://github.com/to-be-specified-upon-acceptance

